# Investigating Biological Aging Measures, Neurocognitive Performance, and Depressive Symptoms: Moderating Effects of HIV and Childhood Maltreatment

**DOI:** 10.64898/2026.11.08.26367522

**Authors:** Kabelo Mabokane Simon Maloka, Georgina Spies, Stefan du Plessis, Sian Megan Joanna Hemmings, Scott Letendre, Soraya Seedat, Jacqueline Samantha Womersley

**Author notes:** Corresponding author: Jacqueline Samantha Womersley Anzio Road Observatory, Cape Town, 7925, South Africa /. E-mail addresses of authors: KMSM GS SDP SMJH SL SS JSW.

## Abstract

**Introduction:** Neurocognitive impairment and depression remain common among people with HIV (PWH), and childhood maltreatment (CM) may further increase vulnerability. In this cohort, we previously reported that PWH exhibited shorter absolute telomere length, lower MRI- based brain-predicted age difference, and greater epigenetic age acceleration (AA) compared to controls, with limited associations with CM severity. Building on this, the present study examines whether these markers are linked to neurocognitive performance and depressive symptoms, and whether HIV status or CM severity moderates such associations.

**Methods:** In this study of 238 women (122 PWH), biological aging was assessed using absolute telomere length, mitochondrial DNA copy number, MRI-based brain-predicted age difference, and DNA methylation AA measures, including Horvath, Hannum, PhenoAge, Horvath intrinsic epigenetic, and Hannum intrinsic epigenetic and extrinsic epigenetic AA. Neurocognitive performance was measured using a standardised assessment battery and global deficit scores (GDS) were derived. Depressive symptoms were assessed with the Centre for Epidemiologic Studies - Depression Scale. Linear regression models examined associations with GDS and depressive symptoms, with interaction models evaluating moderation by HIV status and CM severity.

**Results:** Biological aging markers were not associated with either neurocognitive performance or depressive symptoms. Though HIV infection in combination with higher mitochondrial DNA copy number and extrinsic epigenetic AA was nominally associated with higher GDS (p = 0.012 and p = 0.047, respectively), these associations did not survive correction for multiple testing. Neither HIV nor CM moderated the association between biological aging markers and depression scores.

**Conclusions:** Despite accelerated biological aging being evident in PWH, we found limited evidence of association with neurocognitive performance or depressive symptoms, underscoring their multifactorial nature. Larger longitudinal studies are needed to determine whether these markers predict neurocognitive and psychiatric trajectories.

## Introduction

Neurocognitive impairment and depression are two of the most prevalent and debilitating outcomes affecting people with HIV (PWH).^1,2^ HIV-associated neurocognitive disorders (HAND) affect approximately 50% of PWH globally.^3^ Depression, likewise, affects over 280 million people globally and occurs at rates nearly three times higher in PWH compared to the general population.^4–7^ Both disorders disproportionally affect women, who face unique psychosocial and biological vulnerabilities.^8–10^

Evidence suggests that HIV adversely influences neurocognition and depression through chronic neuroinflammation and direct neurotoxicity.^1,11,12^ Viral persistence in microglia, astrocytes, and memory T cells sustains immune activation, leading to elevated cytokine release, oxidative stress, and synaptic injury.^11,13,14^ This inflammatory milieu disrupts neural circuits involved in cognition and mood regulation, thereby increasing risk for HAND and depression.^1,2^ Childhood maltreatment (CM), encompassing physical, sexual, and emotional abuse as well as physical and emotional neglect, further compounds this risk.^12,15^ Mechanistically, CM induces long-lasting dysregulation of the hypothalamic-pituitary-adrenal (HPA) axis, leading to altered cortisol secretion, heightened stress reactivity, and downstream structural changes in brain regions such as the hippocampus and prefrontal cortex.^16–18^ CM is highly prevalent globally, with particularly elevated rates in sub-Saharan Africa.^19–21^ In South Africa, women are disproportionately affected by both HIV and CM, warranting investigations into how the dual burden of these stressors contributes to neurocognitive and affective outcomes.^19,22–25^

Emerging evidence suggests that biological aging, the progressive decline in physiological function across molecular, cellular, tissue, and organ systems, may serve as a shared pathway linking HIV, CM, neurocognitive impairment, and depression (Bell et al., 2019; Ferrucci et al., 2020). HIV and CM are each recognised as potent stressors that accelerate biological aging, as reflected by markers of biological aging processes such as mitochondrial DNA copy number (mtDNAcn)^26–28^, telomere length (TL)^27,29^, DNA methylation age (DNAmAge) clocks^30,31^, and magnetic resonance imaging-based brain-predicted age difference (MRI-BPAD)^32,33^. These aging indices have, in turn, been linked to poorer neurocognitive performance and elevated depressive symptoms.^34–37^ Collectively, this evidence suggests that accelerated biological aging may represent a mechanistic pathway through which HIV and CM contribute to neurocognitive impairment and depression in affected individuals. However, most prior research has examined these influences largely in isolation, focusing on HIV and CM separately and often relying on a single measure of biological aging.^35,38,39^ Given that HIV-related neuroinflammation and CM- related stress dysregulation are both potent biological stressors, assessing their joint effects is valuable for clarifying whether they converge on shared aging pathways or exert distinct influences. Exploring this intersection can help determine whether dual exposure amplifies vulnerability to neurocognitive and psychiatric outcomes, or whether each factor contributes through independent mechanisms.

This study therefore aims to examine associations between multiple biological aging markers, including absolute TL (aTL), mtDNAcn, MRI-BPAD, and DNAmAge clocks, and two outcomes assessed separately: global deficit scores (GDS) and depressive symptom scores. Given evidence linking HIV and CM to accelerated biological aging and neuropsychiatric vulnerability, we further hypothesised that HIV status and CM severity would modify the associations between biological aging and GDS and depressive symptoms, such that these associations would be stronger among PWH with greater CM severity.

## Methods

### Study design and participants

This cross-sectional analysis drew on baseline data from 238 women enrolled between 2008 and 2015 in a longitudinal study in Cape Town, South Africa (122 PWH). The study received ethics approval from the Health Research Ethics Committee (HREC) of Stellenbosch University, Cape Town, South Africa, and is renewed annually (project ethics code: N07/07/153). All participants provided informed consent, and the study was conducted in accordance with the recommendations of the Declaration of Helsinki. Participants were recruited from community health care facilities and completed sociodemographic and neuropsychological assessments, along with blood collection. Due to data availability, biological aging measures were available for subsets of the cohort i.e., aTL and mtDNAcn for 210 women (105 PWH), MRI-BPAD for 202 (105 PWH), and DNAmAge clocks for 231 (120 PWH). Recruitment procedures have been described previously.^40^ Eligible participants were aged 18-50 years, literate in English/Afrikaans/isiXhosa, medically stable, and psychotropic medication-naïve (including antidepressants). Exclusion criteria included major psychiatric illness, substance use disorder, significant medical comorbidities, prior head injury with over 24 hours loss of consciousness, probable dementia, opportunistic central nervous system (CNS) infection or neoplasm, hepatitis B, or MRI contraindications. HIV infection was determined by enzyme linked immunosorbent assay. For PWH, immunovirological markers were assessed, including CD4+ T cell count (indicator of immune function and disease progression) and CD8+ T cell count (reflecting immune activation), CD4:CD8 ratio (marker of immune balance), and plasma viral load (measure of treatment response and ongoing viral replication).

### Cognitive measures

Neurocognitive performance was assessed using the HIV Neurobehavioral Research Center (HNRC) neuropsychological test^41^, comprising 17 tests across seven domains sensitive to HIV effects: attention/working memory, learning, delayed recall, executive function, motor ability, information processing speed, and verbal fluency.^42^ Raw scores were adjusted using locally derived normative prediction equations based on a sample of 500 South African adults without HIV (Deist et al., 2023). These equations were developed using multiple fractional polynomial modelling and incorporated age, education, ethnicity, and test language.^43–45^ Residuals were converted to demographically corrected T-scores, which were then transformed into deficit scores reflecting impairment severity. Global deficit scores were calculated as averages across all tests and were used continuously in this study. Higher scores indicate poorer neurocognitive performance, whereas lower scores indicate better neurocognitive performance. This approach has been validated in South African cohorts.^45^

### Clinical measures

Depressive symptoms were assessed using the 20-items Center for Epidemiologic Studies Depression Scale (CES-D).^46^ Responses ranged from ‘rarely or none of the time’ to ‘most or all of the time.’ Total scores were calculated by summing item responses and were used as continuous variables in this study. Higher scores indicate more severe depressive symptoms. A commonly used clinical cut-off of ≥ 16 indicates risk for clinically significant depressive symptoms. However, in this study CES-D total scores were analysed as continuous variables. The CES-D has shown excellent reliability in our sample (α = 0.96).

CM was measured with the 28-item Childhood Trauma Questionnaire – Short Form (CTQ-SF), assessing physical, emotional, and sexual abuse, as well as physical and emotional neglect before age 18.^47^ Total scores were analysed continuously, with higher scores indicating greater CM (α = 0.83).

Lifetime trauma exposure was assessed using the Life Events Checklist (LEC), which captures exposure to 17 potentially traumatic events.^48^ The total score comprised the sum of index events directly experienced or witnessed by participants, with higher scores indicating greater trauma exposure (α = 0.77).

### MRI acquisition

Siemens MAGNETOM Allegra 3T scanner was used to acquire high-resolution T1-weighted MRI scans with either a magnetisation-prepared rapid gradient-echo (MPRAGE) sequence (n = 102) (field of view [FOV] = 256 mm; repetition time [TR] = 2080 ms; flip angle = 7°, echo time [TE] = 1.53 ms; voxel size = 1.3 x 1.0 x 1.3 mm; inversion time [TI] = 1100 ms) or a multi-echo MPRAGE sequence (n = 100) (128 slices, TE1 = 1.53 ms, TE2 = 3.21 ms, TE3 = 4.89 ms, TE4 = 6.57 ms, TR = 2530 ms; flip angle = 7°, 1 mm isotropic voxel size, FOV = 256 mm).

### Biological aging measures

Whole-blood DNA concentration and purity were evaluated using a Qubit 4 fluorometer (Invitrogen, Waltham, MA) or NanoDrop One spectrophotometer (Thermo Fisher Scientific, Waltham, MA). Samples were normalised to 350 ng of total DNA and diluted to 6 ng/µL prior to methylation profiling. DNA methylation profiling was performed at the University of Minnesota Genomics Center using the Infinium MethylationEPIC v2 BeadChip (Illumina, San Diego, CA). Quality control and preprocessing were conducted in R using the ENmix pipeline^49^, including assessment and exclusion of low-quality probes and samples based on detection P values, bead counts, bisulfite-conversion intensity, and outlier detection. The resulting beta values were entered into the online DNAmAge calculator (https://dnamage.clockfoundation.org) to estimate Horvath, Hannum, PhenoAge, Horvath intrinsic epigenetic age acceleration (IEAA), Hannum IEAA, and extrinsic epigenetic age acceleration (EEAA). Age acceleration was defined as the difference between estimated DNAmAge and chronological age.

Cellular aging was indexed through aTL and mtDNAcn, which were measured simultaneously in triplicate using the absolute human TL and mtDNAcn dual quantification quantitative polymerase chain reaction (qPCR) assay kit (ScienCell Research Laboratories, Carlsbad, CA; Catalog number 8958) on a QuantStudio 6 Pro Real-Time PCR System (Thermo Fisher Scientific). Samples were analysed in triplicate according to the manufacturer’s protocols. Absolute TL was expressed as kilobases per chromosome end, and mtDNAcn was expressed as mitochondrial DNA copies per diploid cell.

T1-weighted MRI-based brain-predicted age (MRI-BPA) was assessed using the brainageR pipeline.^32^ This model applies Gaussian process regression to voxel-wise tissue maps derived from SPM12 preprocessing and was trained on 3377 healthy adults aged 18 – 92 years and has been validated across multiple cohorts.^50–52^ MRI-BPAD was calculated as MRI-BPA minus chronological age. The brainageR code is openly available (https://github.com/jamescole/brainageR).

Assay and model performance were evaluated using established metrics: correlations with chronological age for all measures, melting curve and reference DNA two-fold serial dilution checks for qPCR assays, and mean absolute error (MAE), root mean squared error (RMSE), and variance explained for brainageR, with separate assessments in PWH and controls.

### Data analysis

Statistical analyses were conducted using R version 4.3.3.^53^ The distribution of continuous variables was assessed using the Shapiro-Wilk test. Demographic and clinical data were summarised using Welch’s t-tests or Mann-Whitney U tests for continuous variables, and chi- square tests for categorical variables. Effect sizes were calculated with Cohen’s d for normally distributed variables and rank-biserial correlation for non-normal variables. Due to extreme values and right-skewed distribution, aTL and mtDNAcn data were log-transformed and winsorised at the 95th percentile.^54,55^ Associations between CTQ-SF scores and aging markers were examined using Spearman correlation. Linear regression models tested the associations between individual aging markers and clinical outcomes, including GDS and CES-D scores. Moderation analyses tested the *a priori* hypothesis that HIV status and CTQ-SF severity would modify the associations between individual aging markers and GDS or CES-D scores. All models were adjusted for age, age-squared (to account for the non-linear association between MRI-BPA and chronological age^56^), MRI protocol type (MRI-BPAD models), plate number (aTL and mtDNAcn models), and LEC scores (CTQ-SF analyses). Multiple testing was controlled using false discovery rate (FDR). Detailed analyses of correlations between biological aging markers and chronological age, group comparisons by HIV status, and associations with CM are reported elsewhere.^57^ In the present study, these findings are briefly summarised to provide context for the outcome analyses.

## Results

### Demographic and clinical characteristics

The analytic sample comprised 238 women with data available for at least one biological aging marker (122 PWH) (see Supplementary Table S1). Compared to controls, PWH were significantly older (U = 4247.0, p < 0.001) and reported lower educational attainment ( *2*(2) = 4.7, *p* = 0.031). Employment status, self-identified ethnicity, and MRI protocol distribution did not differ between groups. CTQ-SF total score and all subtypes were higher among PWH than controls, whereas lifetime traumatic event exposure did not differ significantly. Median GDS was higher in PWH (*U* = 5668.50, *p* = 0.008), while CES-D scores trended higher but did not reach statistical significance (*U* = 6053.00, *p* = 0.051).

### Biological aging markers and chronological age

Comprehensive validation analysis of biological aging markers in this cohort is presented in detail elsewhere.^57^ In brief, log-winsorised aTL and mtDNAcn showed weaker associations with chronological age, MRI-BPA was moderately correlated (r = 0.50, MAE = 5.59 years, RMSE = 7.22 years), and DNAmAge clocks showed strong correlations with chronological age (*r* = 0.77 – 0.83).

### Group comparison of aging markers by HIV status and associations with CM severity

Group comparisons of biological aging markers by HIV status and associations with CM severity are reported in detail elsewhere.^57^ Summarised here, PWH exhibited shorter log-winsorised aTL (*W* = 7947.50, *r_rb_* = 0.38, *p* < 0.001) and higher HorvathAA (Welch *t*(225.8) = -4.45, *d* = −0.59, *p* < 0.001), HannumAA (Welch *t*(229.0) = -2.59, *d* = −0.34, *p* = 0.010), PhenoAA (Welch *t*(228.4) = -6.42, *d* = −0.85, *p* < 0.001), and EEAA (Welch *t*(225.7) = -5.80, *d* = −0.76, *p* < 0.001) compared to controls, while MRI-BPAD was lower in PWH (Welch *t*(199.9) = 2.54, *d* = 0.36, *p* = 0.012). No significant differences were observed for mtDNAcn, Horvath IEAA, or Hannum IEAA. CM severity showed little evidence of association with aging markers after correction for multiple testing.

### Assessing biological aging markers, neurocognitive performance, and depressive symptoms

Linear regression models were conducted to examine associations between biological aging markers and both GDS and CES-D scores (Table 1). For the GDS model, none of the aging markers were significantly associated with overall neurocognitive performance on the cognitive battery. Estimates were small in magnitude (β ranging from –0.012 to 0.003), confidence intervals ranged between −0.121 and 0.106, and all FDR-adjusted *p*-values exceeded 0.278. For the CES-D model, results were similarly null. Most markers showed weak associations (β ranging from –2.474 to 0.218), with wide confidence intervals and non-significant FDR-adjusted *p*-values.

**Table 1.**
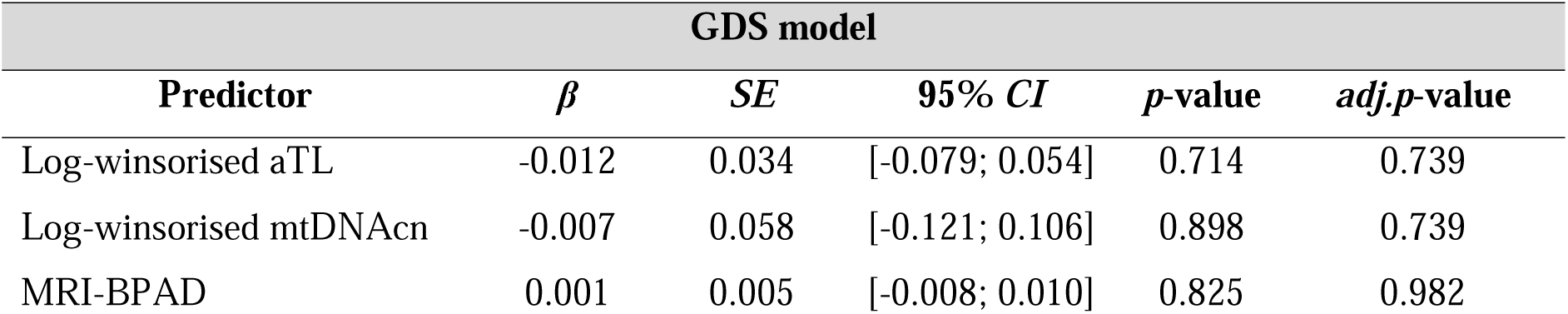
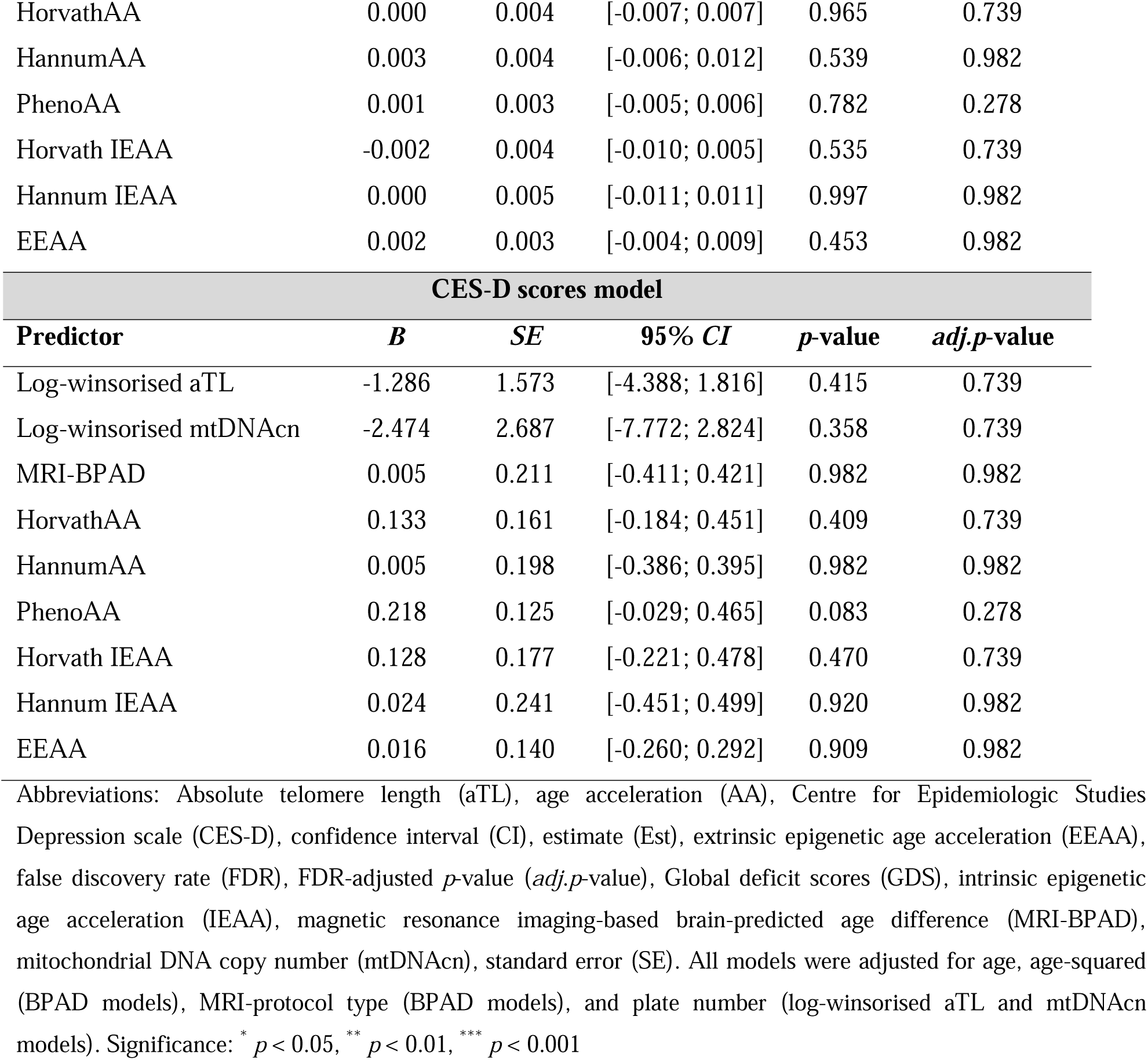
Linear regression models assessing the relationship between biological aging markers and GDS and CES-D scores.

### Moderation analyses by HIV status and childhood maltreatment

Moderation analyses were conducted to examine whether HIV status and CTQ-SF scores modified the associations between biological aging markers and GDS and CES-D scores (Table 2). There was little evidence that HIV status or CM consistently moderated the associations between biological aging markers and GDS. However, nominal associations were observed for the log-winsorised mtDNAcn x HIV interaction (β = 0.880, *p* = 0.012), the log-winsorised mtDNAcn x HIV x CTQ-SF interaction (β = −0.021, *p* = 0.006), the EEAA x HIV interaction (β = 0.046, *p* = 0.047), and the EEAA x HIV x CTQ-SF interaction (β = −0.001, *p* = 0.021).

**Table 2.** Linear regression models testing whether HIV status and CTQ-SF severity moderate the associations between biological aging markers and GDS.

| **GDS model** |  |  |  |  |  |
|---|---|---|---|---|---|
| **Predictor** | ***β*** | ***SE*** | **95% *CI*** | ***p*-value** | ***adj.p*-value** |
| **Log-winsorised mtDNAcn** |  |  |  |  |  |
| Log-win mtDNAcn x HIV | 0.875 | 0.346 | [0.193; 1.557] | 0.012<sup>*</sup> | 0.284 |
| Log-win mtDNAcn x CTQ-SF | 0.009 | 0.006 | [-0.003; 0.022] | 0.142 | 0.544 |
| Log-win mtDNAcn x HIV x CTQ-SF | -0.021 | 0.007 | [-0.036; -0.006] | 0.006<sup>**</sup> | 0.214 |
| **EEAA** |  |  |  |  |  |
| EEAA x HIV | 0.046 | 0.023 | [0.001; 0.092] | 0.047<sup>*</sup> | 0.544 |
| EEAA x CTQ-SF | 0.001 | 0.000 | [0.000; 0.002] | 0.059 | 0.544 |
| EEAA x HIV x CTQ-SF | -0.001 | 0.001 | [-0.002; 0.000] | 0.021<sup>*</sup> | 0.389 |

Abbreviations: Childhood Trauma Questionnaire-Short Form (CTQ-SF), confidence interval (CI), estimate (Est), extrinsic epigenetic age acceleration (EEAA), false discovery rate (FDR), FDR-adjusted *p*-value (*adj.p*-value), global deficit scores (GDS), log-winsorised (Log-win), mitochondrial DNA copy number (mtDNAcn), standard error (SE). All models were adjusted for age, age-squared (MRI-BPAD models), MRI-protocol type (MRI-BPAD models), plate number (log-winsorised aTL and mtDNAcn models), and Life Events Checklist scores (CTQ-SF models). Significance: <sup>\*</sup> *p* < 0.05, <sup>\*\*</sup> *p* < 0.01, <sup>\*\*\*</sup> *p* < 0.001

However, none of these interaction effects remained significant after FDR correction. For CES-D scores, no significant interaction effects for biological aging markers, HIV status, or CTQ-SF scores were observed. The full moderation results are reported in Supplementary Table S2.

## Discussion

The present study examined whether multiple measures of biological aging, including aTL, mtDNAcn, MRI-BPAD, and DNAmAge clocks (HorvathAA, HannumAA, PhenoAA, Horvath IEAA, Hannum IEAA, and EEAA), were associated with overall neurocognitive performance and depressive symptoms, and whether these relationships were moderated by HIV status and CM severity. PWH had shorter aTL, lower MRI-BPAD, greater HorvathAA, HannumAA, PhenoAA, and EEAA than controls. CM was not linked to biological aging markers. Across all models, none of the biological aging markers were significantly associated with neurocognitive performance or depressive symptom severity. Furthermore, there was little evidence that HIV status or CM consistently modified these associations.

The limited relationships between biological aging markers and neurocognitive performance or depressive symptoms suggest that these outcomes are likely influenced by multiple interacting mechanisms, including neuroinflammation, vascular dysfunction, metabolic alterations, psychosocial stress, and neurodegeneration, of which biological aging represents only one component.^58–60^ It is also plausible that accelerated biological aging precedes clinically detectable neuropsychiatric changes, with poor neurocognitive performance or depressive symptoms emerging only after prolonged cumulative biological burden.^61,62^ Longitudinal studies will be critical to determine whether accelerated biological aging predicts decline or symptom onset over time in vulnerable populations.

Our findings differ from several studies reporting associations between accelerated biological aging and poorer global neurocognitive impairment^63,64^ or greater depressive symptom severity^65^, particularly among PWH. However, evidence across the field remains mixed, with some studies reporting null or weak associations with neurocognitive impairment^66,67^ or depression^68,69^. Methodological and cohort differences may partly explain these discrepancies. Prior studies were conducted predominately in high-income countries and mixed-sex cohorts^32,63,65^, whereas our cohort comprised South African women, a population disproportionately affected by both HIV and CM^19,23,24^. Our study cohort was also relatively young, and biological aging effects may be more pronounced in older populations. Sex differences in biological aging and neuropsychiatric vulnerability have been reported^70,71^, raising the possibility that associations between biological aging and neuropsychiatric outcomes vary across populations and contexts.

Measurement considerations may also contribute. While the GDS is a well validated global measure of neurocognitive performance^64,72,73^, it may be less sensitive to subtle domain-specific associations that have been reported in PWH^74,75^. Similarly, the CES-D captures depressive symptom severity but may not fully reflect the complex phenomenology of depression in HIV, which can be influenced by stigma, social adversity, and chronic stress exposure.^10,76,77^ Moreover, studies have reported that DNAmAge clocks may have reduced precision in populations with greater genetic and environmental diversity^78–80^, potentially attenuating associations with functional outcomes. Structural brain aging may also manifest heterogeneously, with fronto-striatal and limbic structures particularly vulnerable to HIV related injury^74,81,82^ and CM stress^83,84^, which could differentially impact cognition and mood. Telomere dynamics may reflect both attrition and compensatory elongation processes, complicating interpretation in cross sectional designs.^85,86^ Similarly, mtDNAcn measured in peripheral blood may not accurately capture mitochondrial function within the CNS, and estimates can be confounded by platelet leukocyte ratios or technical variability in qPCR assays.^87–89^ These considerations underscore the importance of evaluating biological aging across diverse populations, employing both global and domain specific neurocognitive measures, validated depression instruments, and multimodal brain imaging to capture region specific vulnerability.

We found little evidence that HIV status or CM consistently modified the relationships between biological aging markers and neurocognitive performance or depressive symptoms. Although several nominal interactions involving mtDNAcn and EEAA were observed, none remained significant after correction for multiple comparisons, underscoring the limited evidence of robust moderation effects in this cohort. To our knowledge, this is the first study to examine interactions between biological aging markers, HIV, and CM in relation to neurocognitive performance and depressive symptoms. Replication in larger longitudinal cohorts will be important to determine whether subtle interaction effects emerge over time. While moderation effects were not observed, PWH demonstrated accelerated biological aging across several markers, including shorter aTL and greater epigenetic age acceleration, whereas MRI-BPAD was lower compared to controls. These results, reported in detail elsewhere^57^, are consistent with prior evidence linking HIV to telomere shortening and epigenetic age acceleration.^29,30,32,67^ Together, they support the idea that persistent immune activation, neuronal injury, and cellular senescence contribute to accelerated biological aging in HIV.^29,30^ In contrast, CM severity showed little evidence of association with biological aging markers in this cohort, echoing the mixed findings across prior studies.^27,90–92^ These contextual results provide background for interpreting the limited moderation effects observed here and emphasise the importance of future work disentangling shared versus distinct pathways through which HIV and CM may influence biological aging.

This study has several limitations worth noting. The cross-sectional design precludes causal inference, and the relatively small sample size yielded wide confidence intervals, limiting the precision of our estimates of interaction effects.^93^ CM was assessed retrospectively and is therefore susceptible to recall bias.^94^ Neuroimaging analyses were limited to a single modality, and additional modalities such as functional MRI or diffusion tensor imaging may provide more sensitive markers of brain aging.^95,96^ Finally, the high amplification efficiency observed in the mtDNAcn qPCR serial dilution indicates the results should be interpreted cautiously due to potential technical variability.^57^ Despite these limitations, the present study has several strengths, including the use of normative neurocognitive performance scores to reduce sociodemographic bias^44,45^, validation of the brainageR model in a South African cohort, measurement of aTL rather than relative TL to provide a more direct index of telomere dynamics^85^, and a multi- marker approach spanning molecular, cellular, and structural domains of biological aging.

## Conclusions

Accelerated biological aging was evident among PWH but was not associated with neurocognitive performance or depressive symptoms, suggesting that these biological changes alone may be insufficient to explain cognitive and depressive symptomatology. Instead, accelerated biological aging may represent a marker of vulnerability within a broader multifactorial disease process. Larger longitudinal studies are needed to determine whether these markers predict neurocognitive performance or depressive symptoms over time, and whether these trajectories are modified by HIV infection and CM.

## Competing interests

All authors declare that they have no conflicts of interest relevant to the content of this manuscript.

## Authors’ contributions

KMSM: Investigation, formal analysis, data curation, visualisation, writing-original draft, writing-review & editing; GS: supervision, methodology, data collection, writing-review & editing; SD: supervision, methodology, software, writing-review & editing; SMJH: supervision, methodology, writing-review & editing; SL: conceptualisation, funding acquisition, writing- review & editing; SS: conceptualisation, writing-review & editing; JSW: conceptualisation, supervision, methodology, funding acquisition, writing-review & editing.

## Acknowledgements

The authors wish to acknowledge Aqeedah Abbas Roomaney, who assisted with DNA methylation data quality control.

## Funding

Grants awarded to JSW supported this research. This work is based on the research supported in part by the National Research Foundation of South Africa (Grant Number: 137781). The work reported herein was made possible through the SAMRC through its Division of Research Capacity Development under the Early Investigators Programme from funding received from the South African National Treasury. The content hereof is the sole responsibility of the authors and does not necessarily represent the official view of the SAMRC. Research reported in this publication was partly supported by the University of California San Diego Center for AIDS Research International Pilot Grant awarded to JSW and SL (5P30AI036214-23, subaward 705735). Additional research support was provided by a CFAR grant awarded to SS (P30-AI036214) and the HIV Neurobehavioral Research Center (HNRC; National Institutes of Health [grant number P30-MH62512]).

## Data Availability Statement

The datasets presented in this article are not readily available due to ethical and legal restrictions. Requests to access the datasets should be directed to JSW. The authors are open to collaborating and sharing data within the limits of ethical review restrictions and data transfer policies of Stellenbosch University.

## Supporting Information

Supporting Information file 1: Investigating Biological Aging Measures, Neurocognitive Performance, and Depressive Symptoms: Moderating Effects of HIV and Childhood Maltreatment The word document includes 2 tables. Table S1 reports sociodemographic and clinical variables in people with and without HIV and Table S2 reports the full results of HIV and CTQ-SF moderation analyses.

## List of abbreviations

aTL: absolute telomere length
BPA: brain-predicted age
CES-D: Center for Epidemiological Studies – Depression Scale
CI: confidence interval
CM: childhood maltreatment
CNS: central nervous system
CTQ-SF: Childhood Trauma Questionnaire – Short Form
DNAmAge: DNA methylation age
EEAA: extrinsic epigenetic age acceleration
Est: estimate
FDR: false discovery rate
FOV: field of view
GDS: global deficit scores
HAND: HIV-associated neurocognitive disorders
HIV: Human Immunodeficiency Virus
HNRC: HIV Neurobehavioral Research Center
HPA: hypothalamic-pituitary-adrenal axis
IEAA: intrinsic epigenetic age acceleration
LEC: Life Events Checklist
MAE: mean absolute error
RMSE: root mean squared error
MPRAGE: magnetisation-prepared rapid gradient echo
MRI-BPAD: magnetic resonance imaging-based brain-predicted age difference
mtDNAcn: mitochondrial DNA copy number
PWH: people with HIV
qPCR: quantitative polymerase chain reaction
SE: standard error
TE: echo time
TI: inversion time
TL: telomere length

## Notes

### Competing Interest Statement

The authors have declared no competing interest.

### Author Declarations

The Health Research Ethics Committee of Stellenbosch University gave ethical approval for this work.

## References

1. Mudra Rakshasa-Loots A. Depression and HIV: a scoping review in search of neuroimmune biomarkers. Brain Commun. 2023;5(5):fcad231. doi:10.1093/braincomms/fcad231

2. Wei J, Hou J, Su B, et al. The Prevalence of Frascati-Criteria-Based HIV-Associated Neurocognitive Disorder (HAND) in HIV-Infected Adults: A Systematic Review and Meta- Analysis. Front Neurol. 2020;11:581346. doi:10.3389/fneur.2020.581346

3. Zenebe Y, Necho M, Yimam W, Akele B. Worldwide Occurrence of HIV-Associated Neurocognitive Disorders and Its Associated Factors: A Systematic Review and Meta- Analysis. Front Psychiatry. 2022;13:814362. doi:10.3389/fpsyt.2022.814362

4. DeJean D, Giacomini M, Vanstone M, Brundisini F. Patient experiences of depression and anxiety with chronic disease: a systematic review and qualitative meta-synthesis. Ont Health Technol Assess Ser. 2013;13(16):1–33.

5. Necho M, Zenebe Y, Tiruneh C, Ayano G, Yimam B. The Global Landscape of the Burden of Depressive Symptoms/Major Depression in Individuals Living With HIV/AIDs and Its Effect on Antiretroviral Medication Adherence: An Umbrella Review. Front Psychiatry. 2022;13:814360. doi:10.3389/fpsyt.2022.814360

6. Sherr L, Clucas C, Harding R, Sibley E, Catalan J. HIV and Depression – a systematic review of interventions. *Psychology*, Health & Medicine. 2011;16(5):493–527. doi:10.1080/13548506.2011.579990

7. Kuehner C. Why is depression more common among women than among men? Lancet Psychiatry. 2017;4(2):146–158. doi:10.1016/S2215-0366(16)30263-2

8. Rubin LH, Springer G, Martin EM, et al. Elevated Depressive Symptoms Are a Stronger Predictor of Executive Dysfunction in HIV-Infected Women Than in Men. JAIDS Journal of Acquired Immune Deficiency Syndromes. 2019;81(3):274–283. doi:10.1097/QAI.0000000000002029

9. Rubin LH, Neigh GN, Sundermann EE, Xu Y, Scully EP, Maki PM. Sex Differences in Neurocognitive Function in Adults with HIV: Patterns, Predictors, and Mechanisms. Curr Psychiatry Rep. 2019;21(10):94. doi:10.1007/s11920-019-1089-x

10. Spies G, Konkiewitz EC, Seedat S. Incidence and Persistence of Depression Among Women Living with and Without HIV in South Africa: A Longitudinal Study. AIDS Behav. 2018;22(10):3155–3165. doi:10.1007/s10461-018-2072-y

11. Thompson LJP, Genovese J, Hong Z, Singh MV, Singh VB. HIV-Associated Neurocognitive Disorder: A Look into Cellular and Molecular Pathology. IJMS. 2024;25(9):4697. doi:10.3390/ijms25094697

12. Spies G, Ahmed-Leitao F, Fennema-Notestine C, Cherner M, Seedat S. Effects of HIV and childhood trauma on brain morphometry and neurocognitive function. J Neurovirol. 2016;22(2):149–158. doi:10.1007/s13365-015-0379-2

13. Huot N, Bosinger SE, Paiardini M, Reeves RK, Müller-Trutwin M. Lymph Node Cellular and Viral Dynamics in Natural Hosts and Impact for HIV Cure Strategies. Front Immunol. 2018;9:780. doi:10.3389/fimmu.2018.00780

14. Saez-Cirion A, Müller-Trutwin M. The Yellow Brick Road towards HIV Eradication. Trends in Immunology. 2019;40(6):465–467. doi:10.1016/j.it.2019.04.006

15. Wang D, Deng Q, Ross B, et al. Mental health characteristics and their associations with childhood trauma among subgroups of people living with HIV in China. BMC Psychiatry. 2022;22(1):13. doi:10.1186/s12888-021-03658-5

16. Andersen SL, Tomada A, Vincow ES, Valente E, Polcari A, Teicher MH. Preliminary Evidence for Sensitive Periods in the Effect of Childhood Sexual Abuse on Regional Brain Development. JNP. 2008;20(3):292–301. doi:10.1176/jnp.2008.20.3.292

17. Petruccelli K, Davis J, Berman T. Adverse childhood experiences and associated health outcomes: A systematic review and meta-analysis. Child Abuse Negl. 2019;97:104127. doi:10.1016/j.chiabu.2019.104127

18. Rampersaud R, Protsenko E, Yang R, et al. Dimensions of childhood adversity differentially affect biological aging in major depression. Transl Psychiatry. 2022;12(1):431. doi:10.1038/s41398-022-02198-0

19. Titi N, Tomlinson M, Mathews S, et al. Violence and child and adolescent mental health: A whole-of-society response. In: South African Child Gauge 2021/2022. Children’s Institute, University of Cape Town; 2022.

20. Gilbert R, Widom CS, Browne K, Fergusson D, Webb E, Janson S. Burden and consequences of child maltreatment in high-income countries. The Lancet. 2009;373(9657):68–81. doi:10.1016/S0140-6736(08)61706-7

21. Meinck F, Cluver LD, Boyes ME, Loening-Voysey H. Physical, emotional and sexual adolescent abuse victimisation in South Africa: prevalence, incidence, perpetrators and locations. Journal of Epidemiology and Community Health. 2016;70(9):910–916. doi:10.1136/jech-2015-205860

22. Brittain K, Zerbe A, Phillips TK, et al. Impact of adverse childhood experiences on women’s psychosocial and HIV-related outcomes and early child development in their offspring. Global Public Health. 2022;17(11):2779–2791. doi:10.1080/17441692.2021.1986735

23. Steventon Roberts K, Smith C, Toska E, et al. Risk factors for poor mental health among adolescent mothers in South Africa. *Psychology*, Health & Medicine. 2022;27(sup1):67–84. doi:10.1080/13548506.2022.2124295

24. UNAIDS. UNAIDS data 2024: South Africa country factsheet. Published online 2024.

25. Womersley JS, Seedat S, Hemmings SMJ. Childhood maltreatment and HIV-associated neurocognitive disorders share similar pathophysiology: a potential sensitisation mechanism? Metab Brain Dis. 2017;32(5):1717–1733. doi:10.1007/s11011-017-0062-9

26. Ma L, Wang X, Xu H. Mitochondrial DNA damage in HIV infection: a mechanistic driver of immunometabolic dysfunction and chronic inflammation. Front Immunol. 2026;16:1722463. doi:10.3389/fimmu.2025.1722463

27. Ridout KK, Khan M, Ridout SJ. Adverse Childhood Experiences Run Deep: Toxic Early Life Stress, Telomeres, and Mitochondrial DNA Copy Number, the Biological Markers of Cumulative Stress. BioEssays. 2018;40(9):1800077. doi:10.1002/bies.201800077

28. Sun J, Longchamps RJ, Piggott DA, et al. Association Between HIV Infection and Mitochondrial DNA Copy Number in Peripheral Blood: A Population-Based, Prospective Cohort Study. J Infect Dis. 2019;219(8):1285–1293. doi:10.1093/infdis/jiy658

29. Massamba MKY, Madurai L, Archary D, Mahomed S. Telomere length dynamics in adults living with HIV: A systematic review. BMC Infect Dis. 2026;26(1):1062. doi:10.1186/s12879-026-13243-4

30. Bożejko M, Knysz B, Czernicka A, Tarski I, Szymczak A, Małodobra-Mazur M. HIV infection is associated with accelerated epigenetic ageing: A systematic review. Ageing Research Reviews. 2025;112:102884. doi:10.1016/j.arr.2025.102884

31. Green M, Horsfall L, Saied A. A Systematic Review of Adverse Childhood Experiences and Epigenetic Age Acceleration in Later Adult Life Measured With Second and Third Generation Epigenetic Clocks. American J of Med Genetics Pt B. 2026;201(4):270–290. doi:10.1002/ajmg.b.33058

32. Cole JH, Underwood J, Caan MWA, et al. Increased brain-predicted aging in treated HIV disease. Neurology. 2017;88(14):1349–1357. doi:10.1212/WNL.0000000000003790

33. Fleming LL, Ohashi K, Bosquet Enlow M, et al. Sensitive periods for the link between childhood maltreatment and brain aging during adulthood. Neurobiology of Stress. 2025;39:100771. doi:10.1016/j.ynstr.2025.100771

34. Gonzales MM, Garbarino VR, Pollet E, et al. Biological aging processes underlying cognitive decline and neurodegenerative disease. Journal of Clinical Investigation. 2022;132(10):e158453. doi:10.1172/JCI158453

35. Kalungi A, Womersley JS, Kinyanda E, et al. Internalizing Mental Disorders and Accelerated Cellular Aging Among Perinatally HIV-Infected Youth in Uganda. Front Genet. 2019;10:705. doi:10.3389/fgene.2019.00705

36. Shiau S, Arpadi SM, Shen Y, et al. Epigenetic Aging Biomarkers Associated With Cognitive Impairment in Older African American Adults With Human Immunodeficiency Virus (HIV). Clin Infect Dis. 2021;73(11):1982–1991. doi:10.1093/cid/ciab563

37. Zhou A, Wu Z, Zaw Phyo AZ, Torres D, Vishwanath S, Ryan J. Epigenetic aging as a biomarker of dementia and related outcomes: a systematic review. Epigenomics. 2022;14(18):1125–1138. doi:10.2217/epi-2022-0209

38. Malan-Müller S, Hemmings SMJ, Spies G, Kidd M, Fennema-Notestine C, Seedat S. Shorter telomere length - A potential susceptibility factor for HIV-associated neurocognitive impairments in South African women [corrected]. PLoS ONE. 2013;8(3):e58351. doi:10.1371/journal.pone.0058351

39. Womersley JS, Spies G, Tromp G, Seedat S, Hemmings SMJ. Longitudinal telomere length profile does not reflect HIV and childhood trauma impacts on cognitive function in South African women. J Neurovirol. Published online August 26, 2021. doi:10.1007/s13365-021-01009-4

40. Spies G, Fennema-Notestine C, Cherner M, Seedat S. Changes in cognitive function in women with HIV infection and early life stress. AIDS Care. 2017;29(1):14–23. doi:10.1080/09540121.2016.1204417

41. Heaton RK, Cysique LA, Jin H, et al. Neurobehavioral effects of human immunodeficiency virus infection among former plasma donors in rural China. J Neurovirol. 2008;14(6):536–549. doi:10.1080/13550280802378880

42. Rackstraw S. HIV-related neurocognitive impairment – A review. *Psychology*, Health & Medicine. 2011;16(5):548–563. doi:10.1080/13548506.2011.579992

43. Royston P, Altman DG. Regression Using Fractional Polynomials of Continuous Covariates: Parsimonious Parametric Modelling. Applied Statistics. 1994;43(3):429. doi:10.2307/2986270

44. Deist M, Suliman S, Kidd M, et al. Neuropsychological Test Norms for the Assessment of HIV-Associated Neurocognitive Impairment Among South African Adults. AIDS Behav. 2023;27(9):3080–3097. doi:10.1007/s10461-023-04029-9

45. Spies G, Deist M, Suliman S, Seedat S. Screening for HIV-associated neurocognitive impairment: Development and validation of an abbreviated neuropsychological test battery for use in South African clinical settings. Neuropsychology. 2023;37(2):166–180. doi:10.1037/neu0000872

46. Radloff. The CES-D scale: A self-report depression scale for research in the general population. Appl Psychol Meas. 1977;1:385–401.

47. Bernstein DP, Stein JA, Newcomb MD, et al. Development and validation of a brief screening version of the Childhood Trauma Questionnaire. Child Abuse & Neglect. 2003;27(2):169–190. doi:10.1016/S0145-2134(02)00541-0

48. Gray MJ, Litz BT, Hsu JL, Lombardo TW. Psychometric properties of the life events checklist. Assessment. 2004;11(4):330–341. doi:10.1177/1073191104269954

49. Xu Z, Niu L, Taylor JA. The ENmix DNA methylation analysis pipeline for Illumina BeadChip and comparisons with seven other preprocessing pipelines. Clin Epigenet. 2021;13(1):216. doi:10.1186/s13148-021-01207-1

50. Biondo F, Jewell A, Pritchard M, et al. Brain-age is associated with progression to dementia in memory clinic patients. NeuroImage: Clinical. 2022;36:103175. doi:10.1016/j.nicl.2022.103175

51. Boyle R, Jollans L, Rueda-Delgado LM, et al. Brain-predicted age difference score is related to specific cognitive functions: a multi-site replication analysis. Brain Imaging and Behavior. 2021;15(1):327–345. doi:10.1007/s11682-020-00260-3

52. Hendrikse C, Van Den Heuvel LL, Emsley R, Seedat S, Du Plessis S. Increased Brain Age Among Psychiatrically Healthy Adults Exposed to Childhood Trauma. Brain and Behavior. 2025;15(4):e70450. doi:10.1002/brb3.70450

53. R Core Team. R: A language and environment for statistical computing. Published online 2024.

54. Alegría-Torres JA, Velázquez-Villafaña M, López-Gutiérrez JM, et al. Association of Leukocyte Telomere Length and Mitochondrial DNA Copy Number in Children from Salamanca, Mexico. Genetic Testing and Molecular Biomarkers. 2016;20(11):654–659. doi:10.1089/gtmb.2016.0176

55. Chong MR, Narula S, Morton R, et al. Mitochondrial DNA Copy Number as a Marker and Mediator of Stroke Prognosis: Observational and Mendelian Randomization Analyses. Neurology. 2022;98(5). doi:10.1212/WNL.0000000000013165

56. Cole JH, Franke K. Predicting Age Using Neuroimaging: Innovative Brain Ageing Biomarkers. Trends in Neurosciences. 2017;40(12):681–690. doi:10.1016/j.tins.2017.10.001

57. Maloka KMS, Spies G, du Plessis S, et al. Biological aging markers in South African women with HIV and variable exposure to childhood maltreatment. medRxiv. Published online January 1, 2026:2026.10.15.26365601. doi:10.64898/2026.10.15.26365601 [preprint]

58. Blossom V, Ullal SD, D’Souza MM, Ranade AV, Kumar NA, Rai R. Implicating neuroinflammation in hippocampus, prefrontal cortex and amygdala with cognitive deficit: a narrative review. 3 Biotech. 2025;15(9):320. doi:10.1007/s13205-025-04468-2

59. Maes M, Almulla AF, You Z, Zhang Y. Neuroimmune, metabolic and oxidative stress pathways in major depressive disorder. Nat Rev Neurol. 2025;21(9):473–489. doi:10.1038/s41582-025-01116-4

60. Sălcudean A, Bodo CR, Popovici RA, et al. Neuroinflammation—A Crucial Factor in the Pathophysiology of Depression—A Comprehensive Review. Biomolecules. 2025;15(4):502. doi:10.3390/biom15040502

61. Chen Z, Su M, Tu Q, Li J, Wu H. Longitudinal bidirectional association of biological aging acceleration with depressive symptoms in mid-to-late life: evidence from the China Health and Retirement Longitudinal Study. GeroScience. 2025;48(2):3097–3113. doi:10.1007/s11357-025-01845-w

62. Gao X, Geng T, Jiang M, et al. Accelerated biological aging and risk of depression and anxiety: evidence from 424,299 UK Biobank participants. Nat Commun. 2023;14(1):2277. doi:10.1038/s41467-023-38013-7

63. Hulgan T, Kallianpur AR, Guo Y, et al. Peripheral Blood Mitochondrial DNA Copy Number Obtained From Genome-Wide Genotype Data Is Associated With Neurocognitive Impairment in Persons With Chronic HIV Infection. J Acquir Immune Defic Syndr. 2019;80(4):e95–e102. doi:10.1097/QAI.0000000000001930

64. Petersen KJ, Strain J, Cooley S, Vaida F, Ances BM. Machine Learning Quantifies Accelerated White-Matter Aging in Persons With HIV. The Journal of Infectious Diseases. 2022;226(1):49–58. doi:10.1093/infdis/jiac156

65. Mehta SR, Iudicello JE, Lin J, et al. Telomere length is associated with HIV infection, methamphetamine use, inflammation, and comorbid disease risk. Drug and Alcohol Dependence. 2021;221:108639. doi:10.1016/j.drugalcdep.2021.108639

66. Heany SJ, Levine AJ, Lesosky M, et al. Persistent accelerated epigenetic ageing in a longitudinal cohort of vertically infected HIV-positive adolescents. J Neurovirol. 2023;29(3):272–282. doi:10.1007/s13365-023-01130-6

67. Woods SP, Teixeira AL, Martins LB, Fries GR, Colpo GD, Rocha NP. Accelerated epigenetic aging in older adults with HIV disease: associations with serostatus, HIV clinical factors, and health literacy. GeroScience. 2023;45(4):2257–2265. doi:10.1007/s11357-023-00759-9

68. Fletcher BD, Hughes-Waldon G, Marks EJ, et al. Does telomere length mediate the association between early life adversity and mental health in childhood? Journal of Affective Disorders. 2026;394:120600. doi:10.1016/j.jad.2025.120600

69. Martinez RAM, Howard AG, Fernández-Rhodes L, et al. Does biological age mediate the relationship between childhood adversity and depression? Insights from the Detroit Neighborhood Health Study. Social Science & Medicine. 2024;340:116440. doi:10.1016/j.socscimed.2023.116440

70. Merrick R, Brayne C. Sex Differences in Dementia, Cognition, and Health in the Cognitive Function and Ageing Studies (CFAS). JAD. 2024;100(s1):S3–S12. doi:10.3233/JAD-240358

71. Phyo AZZ, Fransquet PD, Wrigglesworth J, Woods RL, Espinoza SE, Ryan J. Sex differences in biological aging and the association with clinical measures in older adults. GeroScience. 2023;46(2):1775–1788. doi:10.1007/s11357-023-00941-z

72. Arentoft A, Troxell K, Alvarez K, et al. HIV Antiretroviral Medication Neuropenetrance and Neurocognitive Outcomes in HIV+ Adults: A Review of the Literature Examining the Central Nervous System Penetration Effectiveness Score. Viruses. 2022;14(6):1151. doi:10.3390/v14061151

73. Van Der Watt ASJ, Spies G, Seedat S. A longitudinal analysis of neurocognitive profiles in South African women with HIV. AIDS Care. 2025;37(3):396–409. doi:10.1080/09540121.2025.2454340

74. Nguchu BA, Zhao J, Lu Y, et al. Human immunodeficiency virus accelerates brain aging and disrupts the trajectory of glymphatic clearance in aging brain. Front Psychiatry. 2025;16:1509093. doi:10.3389/fpsyt.2025.1509093

75. Shiau S, Cantos A, Ramon CV, et al. Epigenetic Age in Young African American Adults With Perinatally Acquired HIV. J Acquir Immune Defic Syndr. 2021;87(4):1102–1109. doi:10.1097/QAI.0000000000002687

76. Armoon B, Fleury MJ, Bayat AH, et al. HIV related stigma associated with social support, alcohol use disorders, depression, anxiety, and suicidal ideation among people living with HIV: a systematic review and meta-analysis. Int J Ment Health Syst. 2022;16(1):17. doi:10.1186/s13033-022-00527-w

77. Glynn TR, Llabre MM, Lee JS, et al. Pathways to Health: an Examination of HIV-Related Stigma, Life Stressors, Depression, and Substance Use. IntJ Behav Med. 2019;26(3):286–296. doi:10.1007/s12529-019-09786-3

78. Cruz-Gonzalez S, Okpala O, Gu E, et al. Methylation clocks fail to generalize across genetically admixed individuals. eLife. 2026;14:RP105343. doi:10.7554/eLife.105343.3

79. Cruz-González S, Okpala O, Gu E, et al. Methylation Clocks Do Not Predict Age or Alzheimer’s Disease Risk Across Genetically Admixed Individuals. Genomics. Preprint posted online October 18, 2024. doi:10.1101/2024.10.16.618588

80. Wu S, Pathak GA, Chen Z. Toward equitable biomarkers of aging: rethinking methylation clocks. Trends in Genetics. 2025;41(7):552–553. doi:10.1016/j.tig.2025.06.001

81. Israel SM, Hassanzadeh Behbahani S, Turkeltaub PE, Moore DJ, Ellis RJ, Jiang X. Different roles of frontal versus striatal atrophy in HIV associated neurocognitive disorders. Human Brain Mapping. 2019;40(10):3010–3026. doi:10.1002/hbm.24577

82. Sanford R, Ances BM, Meyerhoff DJ, et al. Longitudinal Trajectories of Brain Volume and Cortical Thickness in Treated and Untreated Primary Human Immunodeficiency Virus Infection. Clinical Infectious Diseases. 2018;67(11):1697–1704. doi:10.1093/cid/ciy362

83. González-Acosta CA, Rojas-Cerón CA, Buriticá E. Functional Alterations and Cerebral Variations in Humans Exposed to Early Life Stress. Front Public Health. 2021;8:536188. doi:10.3389/fpubh.2020.536188

84. Pollok TM, Kaiser A, Kraaijenvanger EJ, et al. Neurostructural traces of early life adversities: A meta-analysis exploring age- and adversity-specific effects. Neuroscience & Biobehavioral Reviews. 2022;135:104589. doi:10.1016/j.neubiorev.2022.104589

85. Dweck A, Maitra R. The advancement of telomere quantification methods. Mol Biol Rep. 2021;48(7):5621–5627. doi:10.1007/s11033-021-06496-6

86. Ye Q, Apsley AT, Etzel L, et al. Telomere length and chronological age across the human lifespan: A systematic review and meta-analysis of 414 study samples including 743,019 individuals. Ageing Research Reviews. 2023;90:102031. doi:10.1016/j.arr.2023.102031

87. Baccarelli AA, Byun HM. Platelet mitochondrial DNA methylation: a potential new marker of cardiovascular disease. Clin Epigenet. 2015;7(1):44. doi:10.1186/s13148-015-0078-0

88. De Punder K, Entringer S, Martens DS, et al. Whole blood mitochondrial DNA copy number in depressed patients with and without a history of adverse childhood experiences: the role of blood cell composition. The World Journal of Biological Psychiatry. 2026;27(2):162–168. doi:10.1080/15622975.2025.2601134

89. Picard M. Blood mitochondrial DNA copy number: What are we counting? Mitochondrion. 2021;60:1–11. doi:10.1016/j.mito.2021.06.010

90. Colich NL, Rosen ML, Williams ES, McLaughlin KA. Biological aging in childhood and adolescence following experiences of threat and deprivation: A systematic review and meta- analysis. Psychological Bulletin. 2020;146(9):721–764. doi:10.1037/bul0000270

91. Ridout KK, Carpenter LL, Tyrka AR. The Cellular Sequelae of Early Stress: Focus on Aging and Mitochondria. Neuropsychopharmacol. 2016;41(1):388–389. doi:10.1038/npp.2015.301

92. Zhang Z, Ren H, Han R, et al. Impact of childhood maltreatment on aging: a comprehensive Mendelian randomization analysis of multiple age-related biomarkers. Clin Epigenet. 2024;16(1):103. doi:10.1186/s13148-024-01720-z

93. Xu Z, Gao F, Fa A, Qu W, Zhang Z. Statistical power analysis and sample size planning for moderated mediation models. Behav Res. 2024;56(6):6130–6149. doi:10.3758/s13428-024-02342-2

94. Newbury JB, Arseneault L, Moffitt TE, et al. Measuring childhood maltreatment to predict early-adult psychopathology: Comparison of prospective informant-reports and retrospective self-reports. Journal of Psychiatric Research. 2018;96:57–64. doi:10.1016/j.jpsychires.2017.09.020

95. Michael C, Jones NS, Hanson JL, et al. A Systematic Evaluation of the Performance of Multiple Brain Age Algorithms in Two Cohorts of Youth. Human Brain Mapping. 2026;47(2):e70458. doi:10.1002/hbm.70458

96. More S, Antonopoulos G, Hoffstaedter F, Caspers J, Eickhoff SB, Patil KR. Brain-age prediction: A systematic comparison of machine learning workflows. NeuroImage. 2023;270:119947. doi:10.1016/j.neuroimage.2023.119947

