## Supplemental Table for "Investigating Biological Aging Measures, Neurocognitive Performance, and Depressive Symptoms: Moderating Effects of HIV and Childhood Maltreatment"

**Supplementary Material**

**Tables**

| **Table S1.** Comparison of sociodemographic and clinical variables between PWH and controls without HIV | | | | |
| --- | --- | --- | --- | --- |
| **Characteristics** | **Total**  **n = 238** | **PWH**  **n = 122** | **Controls**  **n = 116** | **Statistic** |
| Age, median (IQR) | 29.00 (25.00 – 36.00) | 33.00 (28.00 – 38.00) | 26.00 (22.00 – 33.00) | *W* = 4247.00, *p* < 0.001^***^ |
| Education (≥ grade 9), n (%) | 207 (86.97) | 100 (81.97) | 107 (92.24) | *ꭓ*^2^(2) = 4.67, *p* = 0.031^*^ |
| Employed, n (%) | 70 (29.41) | 40 (32.79) | 30 (25.86) | *ꭓ*^2^(2) = 1.06, *p* = 0.303 |
| Self-identified ethnicity, n (%) |  |  |  | *ꭓ*^2^(2) = 0.21, *p* = 0.650 |
| Black African | 234 (98.32) | 119 (97.54) | 115 (99.14) |  |
| South African Coloured^a^ | 4 (1.68) | 3 (2.46) | 1 (0.86) |  |
| CTQ-SF total score, median (IQR) | 44.00 (31.00 – 58.75) | 52.50 (38.00 – 65.50) | 35.00 (27.75 – 47.00) | *W* = 3749.50, *p* < 0.001^***^ |
| Emotional abuse, median (IQR) | 9.00 (5.00 – 15.00) | 12.00 (6.25 – 18.00) | 8.00 (5.00 – 11.25) | *W* = 5192.50, *p* < 0.001^***^ |
| Emotional neglect, median (IQR) | 10.00 (6.00 – 15.00) | 12.50 (8.00 – 16.00) | 7.00 (5.00 – 11.00) | *W* = 4050.50, *p* < 0.001^***^ |
| Physical abuse, median (IQR) | 5.00 (5.00 – 11.00) | 8.00 (5.00 – 13.75) | 5.00 (5.00 – 6.00) | *W* = 4478.50, *p* < 0.001^***^ |
| Physical neglect, median (IQR) | 9.00 (6.00 – 12.00) | 10.00 (7.25 – 13.75) | 7.00 (5.00 – 9.00) | *W* = 4302.50, *p* < 0.001^***^ |
| Sexual abuse, median (IQR) | 5.00 (5.00 – 7.00) | 5.00 (5.00 – 8.75) | 5.00 (5.00 – 5.00) | *W* = 5540.50, *p* < 0.001^***^ |
| LEC score, median (IQR) | 5.00 (2.25 – 7.00) | 5.00 (3.00 – 7.00) | 4.00 (2.00 – 6.00) | *W* = 6182.50, *p* = 0.091 |
| MRI protocol type, n (%)^b^ |  |  |  | *ꭓ*^2^(2) = 0.00, *p* = 1.000 |
| MPRAGE | 102 (50.50) | 53 (50.50) | 49 (50.50) |  |
| ME-MPRAGE | 100 (49.50) | 52 (49.50) | 48 (49.50) |  |
| GDS, median (IQR) | 0.24 (0.10 – 0.48) | 0.31 (0.10 – 0.57) | 0.17 (0.05 – 0.38) | *W* = 5668.50, *p* = 0.008^**^ |
| CES-D scores, median (IQR) | 5.00 (0.00 – 20.00) | 7.50 (0.00 – 27.00) | 4.00 (0.00 – 15.25)- | *W* = 6053.00, *p* = 0.051 |
| Antiretroviral treatment, n (%) | - | 51 (41.80) | – | – |
| Log_10_ VL, median (IQR) | - | 3.27 (1.64 – 4.31) | – | – |
| CD4 count, median (IQR) | - | 383.00 (252.00 – 591.75) | – | – |
| CD8 count, median (IQR)^c^ | - | 956.00 (710.00 – 1089.25) | – | – |
| CD4:CD8 ratio, median (IQR) | - | 0.47 (0.31 – 0.65) | – | – |
| **Abbreviations:** Centre for Epidemiologic Studies Depression scale (CES-D), Childhood Trauma Questionnaire-Short Form (CTQ-SF), cluster of differentiation 4 (CD4), cluster of differentiation 8 (CD8), global deficit score (GDS), interquartile range (IQR), Life Events Checklist (LEC), magnetization-prepared rapid gradient-echo (MPRAGE), magnetic resonance imaging (MRI), multi-echo magnetization-prepared rapid gradient-echo (ME-MPRAGE), people with HIV (PWH), viral load (VL).  ^a^ “Coloured” is the official census category in South Africa (Statistics South Africa, 2025). The term is used here in its technical demographic sense and is not intended in a derogatory manner.  ^b^ MRI data were only available in a subset of 202 participants.  ^c^ CD8 data were only available for a subset of PWH (n = 44).  **Significance:** ^*^ *p* < 0.05, ^**^ *p* < 0.01, ^***^ *p* < 0.001 | | | | |

| **Table S2.** Linear regression models testing whether HIV status and CTQ-SF scores moderate the associations between biological aging markers and GDS and CES-D scores | | | | | |
| --- | --- | --- | --- | --- | --- |
| **GDS model** | | | | | |
| **Predictor** | ***β*** | ***SE*** | **95% *CI*** | ***p*-value** | ***adj.p*-value** |
| **Log-winsorised aTL** |  |  |  |  |  |
| Log-win aTL x HIV | 0.027 | 0.197 | [-0.362; 0.417] | 0.890 | 0.959 |
| Log-win aTL x CTQ-SF | 0.001 | 0.003 | [-0.005; 0.006] | 0.820 | 0.959 |
| Log-win aTL x HIV x CTQ-SF | 0.002 | 0.004 | [-0.006; 0.009] | 0.699 | 0.879 |
| **Log-winsorised mtDNAcn** |  |  |  |  |  |
| Log-win mtDNAcn x HIV | 0.875 | 0.346 | [0.193; 1.557] | 0.012^*^ | 0.284 |
| Log-win mtDNAcn x CTQ-SF | 0.009 | 0.006 | [-0.003; 0.022] | 0.142 | 0.544 |
| Log-win mtDNAcn x HIV x CTQ-SF | -0.021 | 0.007 | [-0.036; -0.006] | 0.006^**^ | 0.214 |
| **MRI-BPAD** |  |  |  |  |  |
| MRI-BPAD x HIV | 0.001 | 0.022 | [-0.043; 0.044] | 0.968 | 0.968 |
| MRI-BPAD x CTQ-SF | 0.000 | 0.000 | [-0.001; 0.001] | 0.641 | 0.840 |
| MRI-BPAD x HIV x CTQ-SF | 0.000 | 0.000 | [-0.001; 0.001] | 0.867 | 0.959 |
| **HorvathAA** |  |  |  |  |  |
| HorvathAA x HIV | 0.019 | 0.023 | [-0.025; 0.064] | 0.398 | 0.669 |
| HorvathAA x CTQ-SF | 0.000 | 0.000 | [0.000; 0.001] | 0.293 | 0.630 |
| HorvathAA x HIV x CTQ-SF | -0.001 | 0.000 | [-0.002; 0.000] | 0.206 | 0.549 |
| **HannumAA** |  |  |  |  |  |
| HannumAA x HIV | 0.039 | 0.030 | [-0.019; 0.097] | 0.189 | 0.549 |
| HannumAA x CTQ-SF | 0.001 | 0.001 | [0.000; 0.002] | 0.136 | 0.544 |
| HannumAA x HIV x CTQ-SF | -0.001 | 0.001 | [-0.002; 0.000] | 0.098 | 0.544 |
| **PhenoAA** |  |  |  |  |  |
| PhenoAA x HIV | 0.029 | 0.019 | [-0.010; 0.067] | 0.142 | 0.544 |
| PhenoAA x CTQ-SF | 0.000 | 0.000 | [0.000; 0.001] | 0.276 | 0.612 |
| PhenoAA x HIV x CTQ-SF | -0.001 | 0.000 | [-0.002; 0.000] | 0.096 | 0.544 |
| **Horvath IEAA** |  |  |  |  |  |
| Horvath IEAA x HIV | 0.013 | 0.025 | [-0.036; 0.061] | 0.606 | 0.825 |
| Horvath IEAA x CTQ-SF | 0.000 | 0.000 | [-0.001; 0.001] | 0.387 | 0.669 |
| Horvath IEAA x HIV x CTQ-SF | 0.000 | 0.001 | [-0.002; 0.001] | 0.383 | 0.669 |
| **Hannum IEAA** |  |  |  |  |  |
| Hannum IEAA x HIV | -0.006 | 0.034 | [-0.074; 0.062] | 0.866 | 0.959 |
| Hannum IEAA x CTQ-SF | 0.000 | 0.001 | [-0.001; 0.001] | 0.508 | 0.774 |
| Hannum IEAA x HIV x CTQ-SF | 0.000 | 0.001 | [-0.001; 0.001] | 0.913 | 0.959 |
| **EEAA** |  |  |  |  |  |
| EEAA x HIV | 0.046 | 0.023 | [0.001; 0.092] | 0.047^*^ | 0.544 |
| EEAA x CTQ-SF | 0.001 | 0.000 | [0.000; 0.002] | 0.059 | 0.544 |
| EEAA x HIV x CTQ-SF | -0.001 | 0.001 | [-0.002; 0.000] | 0.021^*^ | 0.389 |
| **CES-D scores model** | | | | | |
| **Predictor** | ***Β*** | ***SE*** | **95% *CI*** | ***p*-value** | ***p*-value** |
| **Log-winsorised aTL** |  |  |  |  |  |
| Log-win aTL x HIV | -8.723 | 9.038 | [-26.545; 9.099] | 0.336 | 0.650 |
| Log-win aTL x CTQ-SF | 0.018 | 0.135 | [-0.249; 0.284] | 0.897 | 0.978 |
| Log-win aTL x HIV x CTQ-SF | 0.078 | 0.180 | [-0.277; 0.433] | 0.665 | 0.871 |
| **Log-winsorised mtDNAcn** |  |  |  |  |  |
| Log-win mtDNAcn x HIV | 15.971 | 16.095 | [-15.767; 47.709] | 0.322 | 0.650 |
| Log-win mtDNAcn x CTQ-SF | 0.371 | 0.292 | [-0.205; 0.948] | 0.205 | 0.650 |
| Log-win mtDNAcn x HIV x CTQ-SF | -0.461 | 0.348 | [-1.148; 0.226] | 0.187 | 0.650 |
| **MRI-BPAD** |  |  |  |  |  |
| MRI-BPAD x HIV | -0.152 | 0.992 | [-2.108; 1.804] | 0.878 | 0.972 |
| MRI-BPAD x CTQ-SF | 0.008 | 0.015 | [-0.022; 0.038] | 0.583 | 0.801 |
| MRI-BPAD x HIV x CTQ-SF | -0.011 | 0.019 | [-0.049; 0.027] | 0.556 | 0.801 |
| **HorvathAA** |  |  |  |  |  |
| HorvathAA x HIV | -1.690 | 0.973 | [-3.608; 0.228] | 0.084 | 0.650 |
| HorvathAA x CTQ-SF | -0.009 | 0.017 | [-0.043; 0.025] | 0.591 | 0.801 |
| HorvathAA x HIV x CTQ-SF | 0.038 | 0.021 | [-0.004; 0.080] | 0.079 | 0.650 |
| **HannumAA** |  |  |  |  |  |
| HannumAA x HIV | -1.430 | 1.280 | [-3.952; 1.093] | 0.265 | 0.650 |
| HannumAA x CTQ-SF | 0.007 | 0.022 | [-0.038; 0.051] | 0.768 | 0.905 |
| HannumAA x HIV x CTQ-SF | 0.026 | 0.028 | [-0.029; 0.080] | 0.350 | 0.650 |
| **PhenoAA** |  |  |  |  |  |
| PhenoAA x HIV | -1.258 | 0.837 | [-2.907; 0.391] | 0.134 | 0.650 |
| PhenoAA x CTQ-SF | 0.001 | 0.016 | [-0.031; 0.033] | 0.935 | 0.978 |
| PhenoAA x HIV x CTQ-SF | 0.022 | 0.018 | [-0.014; 0.059] | 0.229 | 0.650 |
| **Horvath IEAA** |  |  |  |  |  |
| Horvath IEAA x HIV | -1.635 | 1.054 | [-3.711; 0.441] | 0.122 | 0.650 |
| Horvath IEAA x CTQ-SF | -0.001 | 0.020 | [-0.040; 0.037] | 0.947 | 0.978 |
| Horvath IEAA x HIV x CTQ-SF | 0.034 | 0.024 | [-0.013; 0.081] | 0.151 | 0.650 |
| **Hannum IEAA** |  |  |  |  |  |
| Hannum IEAA x HIV | -1.776 | 1.481 | [-4.694; 1.142] | 0.232 | 0.650 |
| Hannum IEAA x CTQ-SF | 0.009 | 0.023 | [-0.037; 0.054] | 0.702 | 0.883 |
| Hannum IEAA x HIV x CTQ-SF | 0.032 | 0.030 | [-0.027; 0.092] | 0.282 | 0.650 |
| **EEAA** |  |  |  |  |  |
| EEAA x HIV | -0.688 | 1.013 | [-2.684; 1.308] | 0.498 | 0.771 |
| EEAA x CTQ-SF | 0.005 | 0.020 | [-0.035; 0.045] | 0.824 | 0.946 |
| EEAA x HIV x CTQ-SF | 0.012 | 0.023 | [-0.033; 0.057] | 0.603 | 0.801 |
| **Abbreviations:** Absolute telomere length (aTL), age acceleration (AA), Centre for Epidemiologic Studies Depression scale (CES-D), Childhood Trauma Questionnaire-Short Form (CTQ-SF), confidence interval (CI), estimate (Est), extrinsic epigenetic age acceleration (EEAA), false discovery rate (FDR), FDR-adjusted *p*-value (*adj*.*p*-value), global deficit scores (GDS), intrinsic epigenetic age acceleration (IEAA), log-winsorised (Log-win), magnetic resonance imaging-based brain-predicted age difference (MRI-BPAD), mitochondrial DNA copy number (mtDNAcn), standard error (SE). All models were adjusted for age, age-squared (BPAD only), plate number (log-winsorised aTL and mtDNAcn), and Life Events Checklist scores (CTQ-SF models).  **Significance:** ^*^ *p* < 0.05, ^**^ *p* < 0.01, ^***^ *p* < 0.001 | | | | | |
